# Longitudinal SARS-CoV-2 seroprevalence in a rural and urban community household cohort in South Africa, during the first and second waves July 2020-March 2021

**DOI:** 10.1101/2021.05.26.21257849

**Authors:** Jackie Kleynhans, Stefano Tempia, Nicole Wolter, Anne von Gottberg, Jinal N. Bhiman, Amelia Buys, Jocelyn Moyes, Meredith L. McMorrow, Kathleen Kahn, F. Xavier Gómez-Olivé, Stephen Tollman, Neil A. Martinson, Floidy Wafawanaka, Limakatso Lebina, Jacques du Toit, Waasila Jassat, Mzimasi Neti, Marieke Brauer, Cheryl Cohen, for the PHIRST-C Group

## Abstract

**Background:** SARS-CoV-2 infections may be underestimated due to limited testing access, particularly in sub-Saharan Africa. South Africa experienced two SARS-CoV-2 waves, the second associated with emergence of variant 501Y.V2. In this study, we report longitudinal SARS-CoV-2 seroprevalence in cohorts in two communities in South Africa.

**Methods:** We measured SARS-CoV-2 seroprevalence two monthly in randomly selected household cohorts in a rural and an urban community (July 2020-March 2021). We compared seroprevalence to laboratory-confirmed infections, hospitalisations and deaths reported in the districts to calculate infection-case (ICR), infection-hospitalisation (IHR) and infection-fatality ratio (IFR) in the two waves of infection.

**Findings:** Seroprevalence after the second wave ranged from 18% (95%CrI 10-26%) and 28% (95%CrI 17-41%) in children <5 years to 37% (95%CrI 28-47%) in adults aged 19-34 years and 59% (95%CrI 49-68%) in adults aged 35-59 years in the rural and urban community respectively. Individuals infected in the second wave were more likely to be from the rural site (aOR 4.7, 95%CI 2.9-7.6), and 5-12 years (aOR 2.1, 95%CI 1.1-4.2) or ≥60 years (aOR 2.8, 95%CI 1.1-7.0), compared to 35-59 years. The in-hospital IFR in the urban site was significantly increased in the second wave 0.36% (95%CI 0.28-0.57%) compared to the first wave 0.17% (95%CI 0.15-0.20%). ICR ranged from 3.69% (95%CI 2.59-6.40%) in second wave at urban community, to 5.55% (95%CI 3.40-11.23%) in first wave in rural community.

**Interpretation:** The second wave was associated with a shift in age distribution of cases from individuals aged to 35-59 to individuals at the extremes of age, higher attack rates in the rural community and a higher IFR in the urban community. Approximately 95% of SARS-CoV-2 infections in these two communities were not reported to the national surveillance system, which has implications for contact tracing and infection containment.

**Funding:** US Centers for Disease Control and Prevention

**Research in context:** *Evidence before this study:* Seroprevalence studies provide better estimates of SARS-CoV-2 burden than laboratory-confirmed cases because many infections may be missed due to restricted access to care and testing, or differences in disease severity and health-care seeking behaviour. This underestimation may be amplified in African countries, where testing access may be limited. Seroprevalence data from sub-Saharan Africa are limited, and comparing seroprevalence estimates between countries can be challenging because populations studied and timing of the study relative to country-specific epidemics differs. During the first wave of infections in each country, seroprevalence was estimated at 4% in Kenya and 11% in Zambia. Seroprevalence estimates in South African blood donors is estimated to range between 32% to 63%. South Africa has experienced two waves of infection, with the emergence of the B.1.351/501Y.V2 variant of concern after the first wave. Reported SARS-CoV-2 cases may not be a true reflection of SARS-CoV-2 burden and specifically the differential impact of the first and second waves of infection.

*Added value of this study:* We collected longitudinal blood samples from prospectively followed rural and urban communities, randomly selected, household cohorts in South Africa between July 2020 and March 2021. From 668 and 598 individuals included from the rural and urban communities, respectively, seroprevalence was found to be 7% (95%CrI 5-9%) and 27% (95%CrI 23-31%), after the first wave of infection, and 26% (95%CrI 22-29%) and 41% (95%CrI 37-45%) after the second wave, in rural and urban study districts, respectively. After standardising for age, we estimated that only 5% of SARS-CoV-2 infections were laboratory-confirmed and reported. Infection-hospitalisation ratios in the urban community were higher in the first (2.01%, 95%CI 1.57-2.57%) and second (2.29%, 95%CI 1.63-3.94%) wave than the rural community where there was a 0.75% (95%CI 0.49-1.41%) and 0.66% (95%CI 0.50-0.98%) infection-hospitalisation ratio in the first and second wave, respectively. When comparing the infection fatality ratios for the first and second SARS-CoV-2 waves, at the urban site, the ratios for both in-hospital and excess deaths to cases were significantly higher in the second wave (0.36%, 95%CI 0.28-0.57% in-hospital and 0.51%, 95%CI 0.34-0.93% excess deaths), compared to the first wave in-hospital (0.17%, 95%CI 0.15-0.20%) and excess (0.13%, 95%CI 0.10-0.17%) fatality ratios, p<0.001 and p<0.001, respectively). In the rural community, the point estimates for infection-fatality ratios also increased in the second wave compared to the first wave for in-hospital deaths, 0.13% (95%CI 0.10-0.23%) first wave vs 0.20% (95%CI 0.13%-0.28%) second wave, and excess deaths (0.51%, 95%CI 0.30-1.06% vs 0.70%, 95%CI 0.49-1.12%), although neither change was statistically significant.

*Implications of all the available evidence:* In South Africa, the overall prevalence of SARS-CoV-2 infections is substantially underestimated, resulting in many cases being undiagnosed and without the necessary public health action to isolate and trace contacts to prevent further transmission. There were more infections during the first wave in the urban community, and the second wave in the rural community. Although there were less infections during the second wave in the urban community, the infection-fatality ratios were significantly higher compared to the first wave. The lower infection-hospitalisation ratio and higher excess infection-fatality ratio in the rural community likely reflect differences in access to care or prevalence of risk factors for progression to severe disease in these two communities. In-hospital infection-fatality ratios for both communities during the first wave were comparable with what was experienced during the first wave in India (0.15%) for SARS-CoV-2 confirmed deaths. To our knowledge, these are the first longitudinal seroprevalence data from a sub-Saharan Africa cohort, and provide a more accurate understanding of the pandemic, allowing for serial comparisons of antibody responses in relation to reported laboratory-confirmed SARS-CoV-2 infections within diverse communities.

## Introduction

The first laboratory-confirmed case of coronavirus disease 2019 (COVID-19) in South Africa was announced on March 5, 2020, and the country has since experienced two waves of COVID-19.^1^ A nationwide lockdown from 27 March – 30 April 2020 confined all persons to their homes (excluding essential services), which was followed by a gradual easing of restrictions.^2^ The second wave of infections began in November 2020,^1^ and the country instituted a less restrictive lockdown from 28 December 2020 – 1 March 2021, which prohibited all gatherings with the exception of funerals and mandated a night-time curfew.^2^ Across Africa, the second wave was more severe than the first,^3^ and specifically in South Africa higher weekly incidence, hospitalisations and deaths were reported for the second wave, compared to the first.^4-6^ The second wave in South Africa was coupled with the emergence of a new variant of severe acute respiratory syndrome coronavirus 2 (SARS-CoV-2), 501Y.V2, also known as B.1.351.^7^

South Africa reported more than 1.6 million laboratory-confirmed cases (by reverse transcription polymerase chain reaction (RT-PCR) or rapid antigen tests) by mid-May 2021,^4^ but many cases may go undiagnosed due to mild or absent symptoms or lack of, or reluctance to access care or testing. Data on the proportion of people with serologic evidence of prior SARS-CoV-2 infection are critical to assess infection rates, calculate infection-hospitalisation and infection-fatality ratios, compare infection burden between waves of infection and to guide public health responses.^8^ Previous studies have shown that SARS-CoV-2 seroprevalence is higher in close contacts of cases and high-risk healthcare-workers, and lower in individuals younger than 20 years, or 65 years and older, with no differences between males and females.^9^ It is still unclear whether HIV-infection increases the risk for SARS-CoV-2 infection, and results from studies thus far have varied.^10,11^

We describe the seroprevalence of SARS-CoV-2 by age and HIV-infection status in two household cohorts in a rural and an urban community at five time points from July 2020, during the first wave, to March 2021, after the second epidemic wave. We also compare disease burden between the first and second wave by comparing the seroprevalence by wave to reported laboratory-confirmed infections, hospitalisations and deaths within the respective districts in which these two communities are located.

## Methods

### Study population

We conducted a prospective study on a randomly selected household cohort in a rural (Agincourt, Ehlanzeni District, Mpumalanga Province) and an urban (Jouberton, Dr Kenneth Kaunda District, North West Province) community as part of the PHIRST-C study (a **P**rospective **H**ousehold study of SARS-**C**oV-2, **I**nfluenza, and **R**espiratory **S**yncytial virus community burden, **T**ransmission dynamics and viral interaction in South Africa). Methods for the cohort study are detailed in the appendix. Recruitment to this study began in July 2020 and follow-up will continue through July 2021. Households which previously participated in the PHIRST study (Prospective Household observational cohort study of Influenza, Respiratory Syncytial virus and other respiratory pathogens community burden and Transmission dynamics in South Africa) during 2016-2018,^12,13^ and additional randomly selected households were eligible. Households with three or more household members of any age were enrolled if ≥80% of members consented.

The study was approved by the University of the Witwatersrand Human Research Ethics Committee (Reference 150808) and the US Centers for Disease Control and Prevention relied on local clearance (IRB #6840).

### Seroprevalence

We collected baseline data, including demographics and HIV status, and blood (blood draw 1) at enrolment (20 July – 17 September 2020), and every two months thereafter (blood draw 2 during 21 September – 10 October; blood draw 3 during 23 November – 12 December 2020; blood draw 4 during 25 January – 20 February 2021 and blood draw 5 during 22 March – 11 April 2021). HIV status was confirmed from medical records (if HIV-infected), and by rapid test for participants with unknown, or self-reported negative status. Prior SARS-CoV-2 infection was determined using the Roche Elecsys® Anti-SARS-CoV-2 assay (Roche Diagnostics, Rotkreuz, Switzerland), using recombinant nucleocapsid (N) protein. The assay was performed on the Cobas e601 instrument, and a cut-off index (COI) of ≥1.0 was considered an indication of prior infection (seropositivity). Signal to cut-off ratio was not considered. Data analysis was performed in Stata 14 (StataCorp, College Station, Texas, USA), using six age groups: pre-school (<5 years), primary school (5-12 years), secondary school (13-18 years), young adults (19-34 years), adults (35-59 years) and older adults (≥60 years) who are prioritised for SARS-CoV-2 vaccination.^14^ Seroprevalence estimates were adjusted for sensitivity and specificity as previous described,^15^ based on the manufacturers’ reported 99.5% sensitivity and 99.8% specificity.^16^ Seroprevalence 95% credible intervals (95%CrI) were obtained using Bayesian inference with 10,000 posterior draws.^15^ Pearson’s chi-squared test was used to assess the statistical significance of differences in SARS-CoV-2 seropositivity across blood collection times, waves of infection and between the two communities.

### Calculation of infection-case ratio (ICR), infection-hospitalisation ratio (IHR) and infection-fatality ratio (IFR) by wave of infection

To assess the burden of SARS-CoV-2, and compare the severity of illness between the first and second waves, we performed an ecological study comparing estimated number of infections based on seroprevalence in our cohort study, to reported number of cases, hospitalisations and in-hospital and excess deaths in the same district for each wave. We calculated the age-adjusted total number of infections, laboratory-confirmed cases, hospitalisations, deaths, ICR (number of infections compared to laboratory-confirmed cases), IHR and in-hospital and excess death IFR for each wave of infection as described in below equations. The first wave was defined as 1 March 2020 (week 11) to 21 November 2020 (week 47) coinciding with the first case of SARS-CoV-2 reported in South Africa, and ending the week before blood draw 3 started, and the second wave as 22 November 2020 (week 48) to 27 March 2021 (week 11), starting directly after the defined wave 1 period, and ending the week before blood draw 5 started (Figure 1). Data sources used in these calculations are described in the appendix. In brief, all laboratory-confirmed SARS-CoV-2 infections in South Africa (by RT-PCR and antigen tests) are reported to the Notifiable Medical Conditions Surveillance System (NMCSS) through automated feeding of data from public and private laboratory information systems to a data warehouse.^4^ The number of reported laboratory-confirmed SARS-CoV-2 cases from the two districts where the rural and urban communities were located were obtained from the NMCSS,^4^ and numbers of hospitalisations and in-hospital deaths were obtained from the COVID-19 National Hospital Surveillance (DATCOV).^5^ Provincial excess deaths per 100,000 population (based on death trends for 2014-2019) were obtained from the South African Medical Research Council report on weekly deaths.^6^ District-level and South African population denominators were obtained from the StatsSA mid-year population estimates for 2020.^17^ Age standardised estimates for the selected endpoints for each wave were obtained as follows:

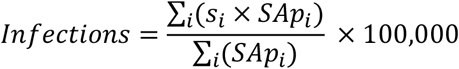

**Figure 1.**
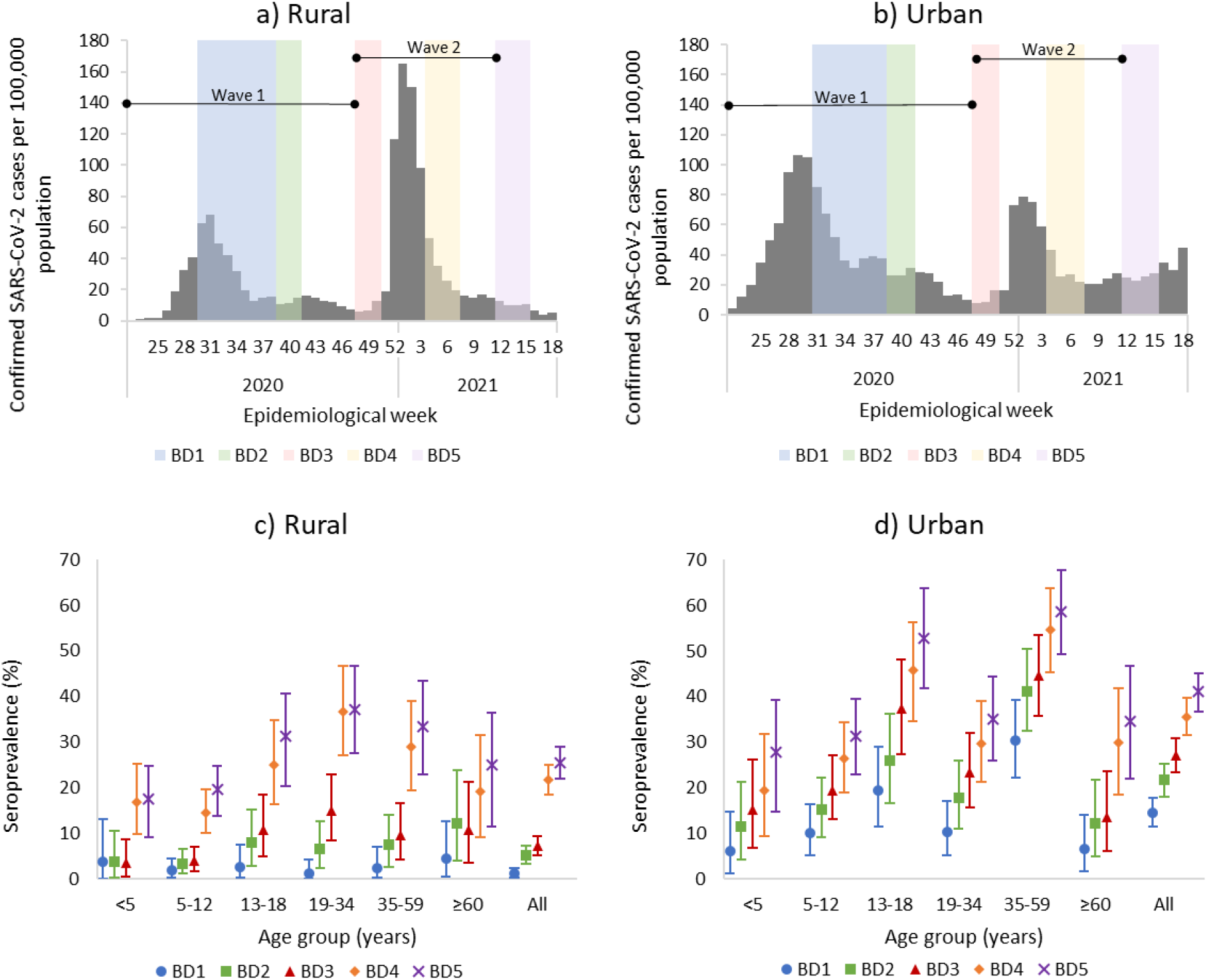
Timing of blood collection and district SARS-CoV-2 weekly incidence^1^ in a) rural community and b) urban community, and seroprevalence at each blood collection by age group in the c) rural and d) urban community, March 2020 – March 2021, South Africa. Blood draw (BD) 1: 20 July – 17 September 2020, blood draw 2: 21 September – 10 October 2020, blood draw 3: 23 November – 12 December 2020, blood draw 4 25 January – 20 February 2021, blood draw 5: 22 March – 11 March 2021. Vertical lines represent 95% credible interval. Seroprevalence estimates adjusted for sensitivity and specificity of assay. Wave 1 (1 March – 21 November 2020) and 2 (22 November 2020 – 27 March 2021) lines indicate period used for analysis. ^1^Laboratory-confirmed SARS-CoV-2 infections (reverse transcription polymerase chain reaction and antigen tests) reported to the Notifiable Medical Conditions Surveillance System (NMCSS).

Where *S*_*i*_ is the seroprevalence in the cohort in the respective community and wave for age group *i* and *P*_*i(SA)*._ is the South African population for age group *i*. Calculated for wave 1 as seroprevalence at blood draw 3, and for wave 2 as seroprevalence at blood draw 5, excluding those who seroconverted at blood draw 3. Estimates only included participants with a blood draw 3 and 5 pair, and adjusted for sensitivity and specificity of test.^15^

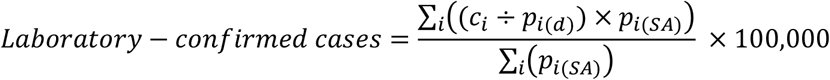

Where *c*_*i*_ is the number of laboratory-confirmed cases (RT-PCR and antigen-based tests) from the respective district reported to the NMCSS (wave 1: 3 March – 21 November 2020, wave 2: 22 November 2020 – 27 March 2021) in age group *i, P*_*i(d)*_ is the district population for age group *i* and *P*_*i(SA)*_ is the South African population for age group *i*.

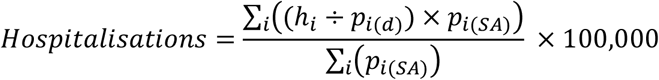

Where *h*_*i*_ is the number of hospitalisations from the respective district reported to COVID-19 Sentinel Hospital Surveillance (DATCOV, wave 1: 5 March – 21 November 2020, wave 2: 22 November 2020 – 27 March 2021)^5^ in age group *i, p*_*i(d)*_ is the district population for age group *i* and *p*_*i(SA)*_ is the South African population for age group *<u>i</u>*

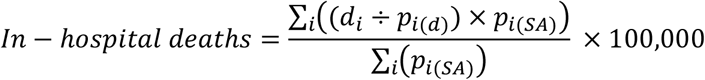

Where *d*_*i*_ is the number of in-hospital deaths from the respective districts reported to DATCOV (wave 1: 5 March – 21 November 2020, wave 2: 22 November 2020 – 27 March 2021)^5^ in age group *i, p*_*i(d)*_ is the district population for age group *i* and *p*_*i(SA)*_ is the South African population for age group *i*.

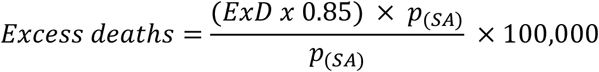

Where *EXD* is the rate of provincial excess deaths adjusted to the South African population reported by South African Medical Research Council (rural wave 1: 28 June – 21 November 2020, urban wave 1: 21 June – 21 November 2020, both communities wave 2: 22 November 2021 – 26 March 2021),^6^ and *p*_*(SA)*_ is the total South African population. According to estimates only 85% of excess deaths are attributable to COVID-19.^6^

For infections, hospitalisations, in-hospital (minimum) and excess (maximum) deaths, 95% confidence intervals (95% CI) were calculated using the Clopper-Pearson method with a Poisson distribution.

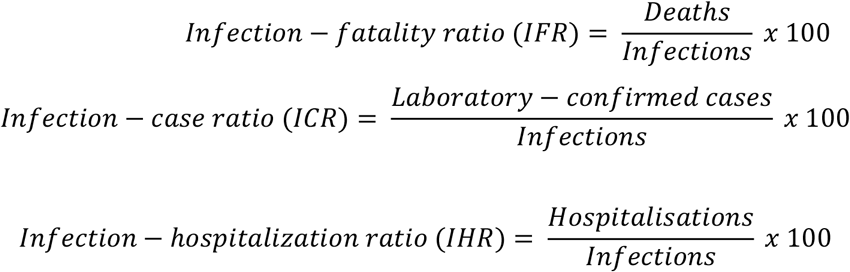

Confidence intervals for infection ratios were calculated as ratios from the 95% confidence intervals of infection, hospitalisation and death rates. For international comparisons, we repeated the calculations to standardise to the WHO standard world population,^18^ using Sprague multipliers^19^ to expand age groups to one-year bands, and aggregate to the age groups used in this study.

### Comparison of cases between first and second wave of infection

We further compared the characteristics of participants who seroconverted during the first wave of infections, to those who seroconverted during the second wave of infections using unconditional logistic regression. We compared site, age, sex, HIV status, CD4 count, viral load, other underlying medial conditions, body mass index, employment status, smoking and alcohol use in participants that seroconverted in wave 1 (blood draw 3) to those who seroconverted in wave 2 (blood draw 5, excluding draw 3 seroconversions). For this analysis, we only included participants with a blood 3 and 5 paired serum sample. For the multivariable model we assessed all variables that were significant at p < 0.2 on univariate analysis, and dropped non-significant factors (p ≥ 0.05) with manual backward elimination.

### Persistence of SARS-CoV-2 antibodies

For all participants for which 5 sera were collected, and who seroconverted during draw 2 to 5, we plotted COI values with the blood draw at which seroconversion took place as point 0. For those who were seropositive at baseline we plotted the COI results from each blood draw. We calculated the mean COI and exact 95% confidence interval at each point using the Clopper-Pearson method. We assessed the percentage of participants with a COI ≥1.0 at each subsequent blood draw as the number of participants with COI ≥1 divided by the seroconverted participants with a serum sample at the time point.

## Results

### Study population

In the rural community, we approached 185 households, 118 (64%) were enrolled and 641/692 (92%) of household members consented and/or assented to participate. In the urban community, 352 households were approached, 114 (32%) enrolled and 570/607 (93%) of household members consented and/or assented. In both communities, the percentage of children, females, and unemployed individuals included in the cohort were higher than in district census data (Appendix Table 1). Median age was 13 (IQR 7-29) and 21 (IQR 10-43) years, and HIV prevalence was 14% (95%CI 11-17%) and 18% (95%CI 14-21%) in the rural and urban communities, respectively.

**Table 1.** Comparison of participants with detectable SARS-CoV-2 antibodies after the first wave (blood draw 3) and second wave (blood draw 5), July 2020 – April 2021, South Africa.

|  | Infected in wave 2 (n/N %) | Univariate OR | 95% CI | Multivariable aOR | 95% CI |
| --- | --- | --- | --- | --- | --- |
| Site |  |  |  |  |  |
| Rural | 100/140 (71) | **4.9** | **3.1-7.8** | **4.7** | **2.9-7.6** |
| Urban | 71/210 (34) | Ref |  | Ref |  |
| Sex |  |  |  |  |  |
| Male | 58/123 (47) | Ref |  |  |  |
| Female | 113/227 (50) | 1.1 | 0.7-1.7 |  |  |
| Age group (years) |  |  |  |  |  |
| <5 years | 16/25 (64) | **3.2** | **1.3-8.0** | 2.7 | 1.0-7.2 |
| 5-12 years | 44/74 (59) | **2.6** | **1.4-4.9** | **2.1** | **1.1-4.2** |
| 13-18 years | 28/64 (44) | 1.4 | 0.7-2.7 | 1.3 | 0.6-2.6 |
| 19-34 years | 33/67 (49) | 1.7 | 0.9-3.3 | 1.3 | 0.7-2.6 |
| 35-59 years | 33/92 (36) | Ref |  | Ref |  |
| ≥60 years | 17/28 (61) | **2.8** | **1.2-6.6** | **2.8** | **1.1-7.0** |
| HIV status |  |  |  |  |  |
| Negative | 132/271 (49) | 1.0 | 0.6-1.7 |  |  |
| Positive | 33/68 (49) | Ref |  |  |  |
| CD4 count |  |  |  |  |  |
| ≥200 | 26/54 (48) | 1.9 | 0.2-21.7 |  |  |
| <200 | 1/3 (33) | Ref |  |  |  |
| Viral load |  |  |  |  |  |
| <1000 | 23/51 (45) | Ref |  |  |  |
| ≥1000 | 5/7 (71) | 3.0 | 0.5-17.2 |  |  |
| Other underlying illness* |  |  |  |  |  |
| No | 155/316 (49) | 1.1 | 0.5-2.2 |  |  |
| Yes | 16/34 (47) | Ref |  |  |  |
| Body-mass index |  |  |  |  |  |
| Underweight | 12/22 (55) | 1.5 | 0.6-3.9 |  |  |
| Normal weight | 76/141 (54) | 1.5 | 0.9-2.5 |  |  |
| Overweight | 38/84 (45) | 1.1 | 0.6-1.9 |  |  |
| Obese | 45/103 (44) | Ref |  |  |  |
| Currently smoking† |  |  |  |  |  |
| No | 81/190 (43) | Ref |  |  |  |
| Yes | 16/36 (44) | 1.1 | 0.5-2.2 |  |  |
| Alcohol use† |  |  |  |  |  |
| No | 79/167 (47) | 2.0 | 1.1-3.8 |  |  |
| Yes | 18/59 (31) | Ref |  |  |  |
| Employment status‡ |  |  |  |  |  |
| Unemployed | 58/128 (45) | 1.9 | 0.5-6.4 |  |  |
| Student | 4/13 (31) | Ref |  |  |  |
| Employed | 16/41 (39) | 1.4 | 0.4-5.5 |  |  |

Includes all participants with blood draw 3 and 5 sera pairs, and who had seroconverted at either draw. \* Self-reported history of asthma, lung disease, heart disease, stroke, spinal cord injury, epilepsy, organ transplant, immunosuppressive therapy, organ transplantation, cancer, liver disease, renal disease, or diabetes. † Individuals ≥15 years. ‡ Individuals ≥18 years.

### Seroprevalence

During blood draw 1-5, blood draw coverage was 67% - 88% and 84-88% in the rural and urban communities, respectively (Appendix Table 2). The majority, 83% (n=553) of participants who lived in the rural and 83% (n=499) in the urban community had both blood draw 3 and blood draw 5 blood collected, with 56% (n=377) and 72% (n=431) of participants at the rural and urban site having all 5 bloods collected, respectively.

Seroprevalence, adjusted for assay sensitivity and specificity, in the rural community was lower at blood draw 1 than in the urban community (1%, 95%CI 0-2% vs 15%, 95%CI 12-18%, p<0.001), increasing after the first wave of infections (at blood draw 3) to 7% (95%CI 5-9) in the rural community and 27% (23-31%) in the urban community (p<0.001, Figure 1, Appendix Table 3). After the second wave (blood draw 5), seroprevalence increased by 19% to reach 26% (95%CI 22-29%, p<0.001) in the rural community, and by 14% to reach 41% (95%CI 37-45%, p<0.001) in the urban community (Figure 1, Appendix Table 3).

At blood draw 5, seroprevalence was highest in the 19-34 years age group (37%, 95% CrI 28-47%) in the rural community and the 35-59 years age group (59%, 95%CrI 49-68%) in the urban community (Figure 1, Appendix Table 3). The seroprevalence was lowest in children <5 years, 18% (95% CrI 10-26%) and 28% (95% CrI 17-41%) in the rural and urban communities, respectively.

At blood draw 5, SARS-CoV-2 seroprevalence was similar between HIV-infected and HIV-uninfected participants in all age groups and in both communities (Appendix Table 4). For adults aged 19-34 years, the seroprevalence in HIV-uninfected individuals in the rural and urban community was 34% (21/66) and 36% (26/73), respectively, compared to HIV-infected individuals at 43% (10/23) and 37% (7/19) in rural and urban community, respectively (p=0.41 and p=0.92, Appendix Table 4).

### Infection-case ratio (ICR), infection-hospitalisation ratio (IHR) and infection-fatality ratio (IFR) by district and wave of infection

Weekly incidence of reported laboratory-confirmed SARS-CoV-2 infections in the rural site district peaked in the first wave at 68 cases per 100,000 population in week 31 of 2020, (starting 26 July), and again at 165 cases per 100,000 in week 2 of 2021 (starting 10 January). In the urban site district, the first wave peaked at 106 cases per 100,000 in week 30 of 2020 (starting 19 July), the second wave at 79 cases per 100,000 in week 2 of 2021 (starting 10 January, Figure 1).

During the first wave of infections (blood draw 3) in the rural community, 40/553 participants had seroconverted resulting in an age-adjusted seroprevalence of 10% (95%CrI 5-17%) or incidence of 10,041 (95%CrI 4,759-17,088) per 100,000 population. Within the rural district, standardised to the South African population, 557 laboratory-confirmed SARS-CoV-2 infections per 100,000 population,^4^ 75 COVID-19-related hospitalisations per 100,000 population, and 14 in-hospital deaths per 100,000 population^5^ were reported by end of week 47 of 2020 (starting 15 November). Excess deaths of 51 per 100,000 population were reported for Mpumalanga Province.^6^ Considering the 10% seroprevalence at blood draw 3 in the rural community as a proxy for the district, the ICR was only 5.55% (95%CI 3.40-11.23%). There was a 0.75% (95%CI 0.49-1.41%) IHR and an in-hospital IFR of 0.13% (95%CI 0.10-0.23%) and an excess deaths IFR of 0.51% (95%CI 0.30-1.06%, Figure 2-3).

**Figure 2.**
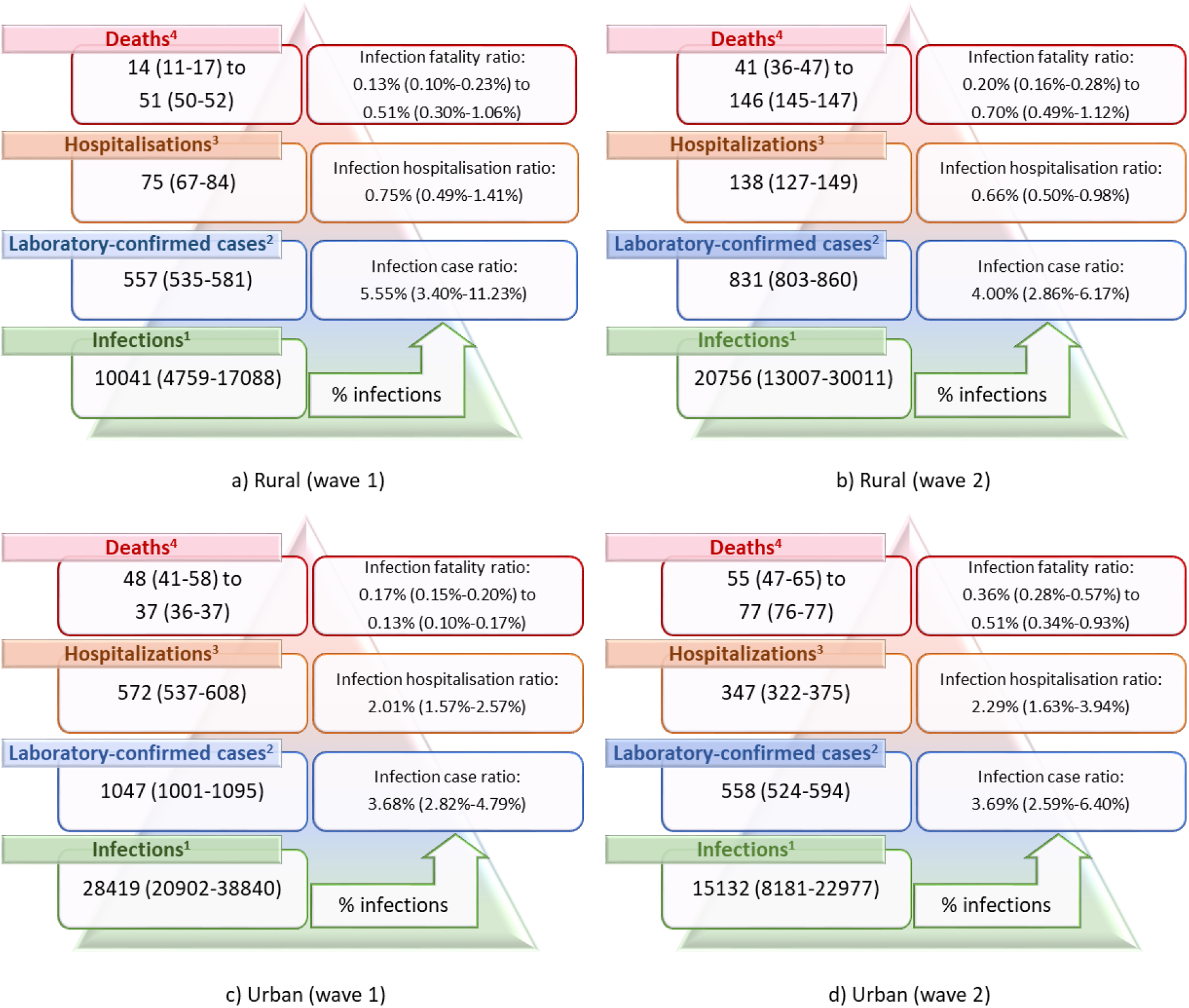
South African age-standardised SARS-CoV-2 infection, diagnosis, hospitalisation and deaths per 100,000 population in a rural community during a) wave 1 and b) wave 2, and urban community during c) wave 1 and d) wave 2 of infections, March 2020 - March 2021, South Africa. ^1^Based on seroprevalence at third/fifth blood draw. ^2^Laboratory confirmed SARS-CoV-2 cases reported from districts from national SARS-CoV-2 surveillance (NMCSS, wave 1: 3 March – 21 November 2020, wave 2: 22 November 2020 – 27 March 2021). ^3^Hospitalisations in the district based on COVID-19 Sentinel Hospital Surveillance (DATCOV, wave 1: 5 March – 21 November 2020, wave 2: 22 November 2020 – 27 March 2021). ^4^Minimum estimate: in-hospitalisation deaths in districts based on COVID-19 Sentinel Hospital Surveillance report (DATCOV, wave 1: 5 March – 21 November 2020, wave 2: 22 November 2020 – 27 March 2021), maximum estimate: provincial excess deaths reported by South African Medical Research Council (rural wave 1: 28 June – 21 November 2020, urban wave 1: 21 June – 21 November 2020, wave 2: 22 November 2021 – 26 March 2021). ^5^Standardised to South Africa mid-year population estimate for 2020. Wave 1: 1 March – 21 November 2020), wave 2 22 November 2020 – 27 March 2021. Values in bracket refer to 95% credible interval for infections and 95% confidence interval for all other estimates.

**Figure 3.**
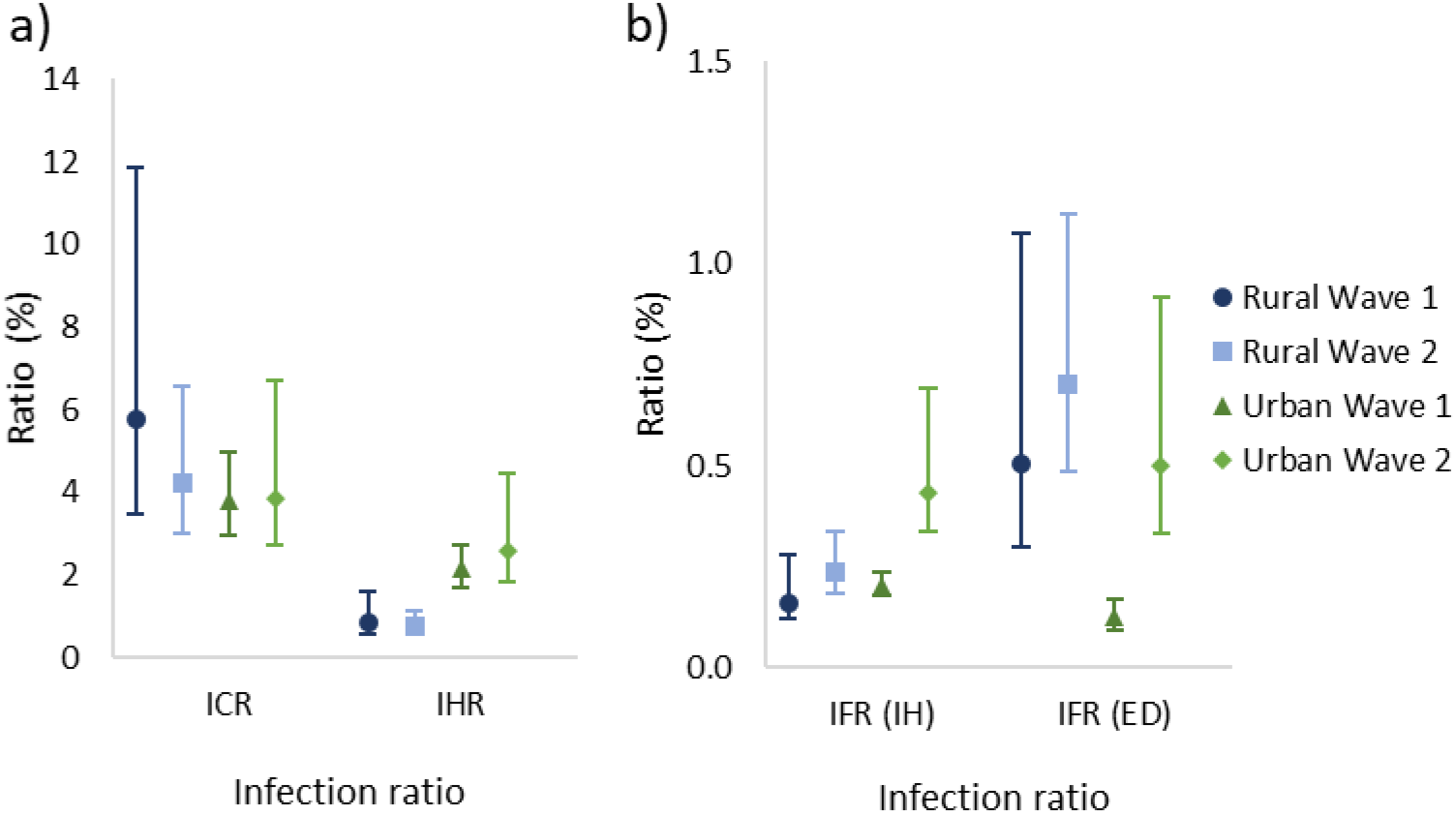
SARS-CoV-2 a) infection-case (ICR), infection-hospitalisation (IHR) and b) in-hospital (IH) infection-fatality (IFR) and excess (ED) infection-fatality (IFR) ratios in a rural and urban community during the first and second wave of infections, March 2020 - March 2021, South Africa. Vertical lines represent 95% confidence interval. Wave 1: 1 March – 21 November 2020, wave 2: 22 November 2020 – 27 March 2021.

If excluding participants who had seroconverted after wave 1, there were 100/553 participants from the rural community that had seroconverted at blood draw 5, an age-adjusted seroprevalence of 21% (95%CrI 13-30%) or incidence of 20,756 (95%CrI 13,007-30,011) per 100,000 population for the second wave. The ICR was 4.00% (95%CI 2.86-6.17%), IHR 0.66% (0.50-0.98%), in-hospital IFR 0.20% (95%CI 0.16-0.28%) and excess deaths IFR 0.70% (95%CI 0.49-1.12%, Figure 2-3).

In the urban community the age-adjusted seroprevalence at blood draw 3 was 29% (95%CrI 21-40%) or incidence of 28,419 (95%CrI 20,902-38,840) per 100,000 population. There was a 3.68% (95%CI 2.82-4.79%) ICR and 2.01% (95%CI 1.57-2.57%) IHR. The in-hospital IFR was 0.17% (95%CI 0.15-0.20%) and excess deaths IFR 0.13% (95%CI 0.10-0.17%, Figure 2-3). During the second wave, the age-adjusted seroprevalence in the urban community was 15% (95%CrI 8%-23%) or incidence 15,132 (95%CrI 8,181-22,977) per 100,000 population, resulting in a ICR estimate of 3.69% (95%CI 2.59-6.40%), IHR of 2.29% (95%CI 1.63-3.94%), in-hospital IFR of 0.36% (95%CI 0.28-0.57%) and an excess deaths IHR of 0.51% (95%CI 0.34-0.93%, Figure 2-3). These estimates standardised to the WHO world population are presented in Appendix Figure 2.

### Comparison of cases between first and second wave of infection

Compared to the urban community, individuals in the rural community who seroconverted were 4.7 (95%CI 2.9-7.6) times more likely to seroconvert during the second wave. Compared to those aged 35-59 years, individuals aged 5-12 years and ≥60 years were 2.1 (95%CI 1.1-4.2) and 2.8 (95%CI 1.1-7.0) times more likely to seroconvert in the second wave (Table 1).

### Persistence of SARS-CoV-2 antibodies

Of the 72 participants seropositive at the baseline blood collection, and with blood draw 1-5 samples collected, 99% (71/72) still had a COI ≥1 by blood draw 5. The mean COI at baseline for seropositive participants was 64, which increased to 125 at blood draw 2 and reduced to 59 at blood draw 5 (Figure 4a). The participant who no longer had detectable SARS-CoV-2 antibodies at the fifth draw had a starting COI of 9, which subsequently declined to 0.7 six months after the first draw. Of the 210 participants with blood draw 1-5 samples, 99% (167/169), 99% (70/71%) and 93% (41/44) still had a COI ≥1 in the first, second and third blood draw after initial seroconversion, respectively (Figure 4b). The participants who seroreverted had starting COIs ranging from 2 – 6. The mean COI at the point of seroconversion was 48, which increased to 86 at the first blood post seroconversion and reduced to 61 at the third draw post seroconversion.

**Figure 4.**
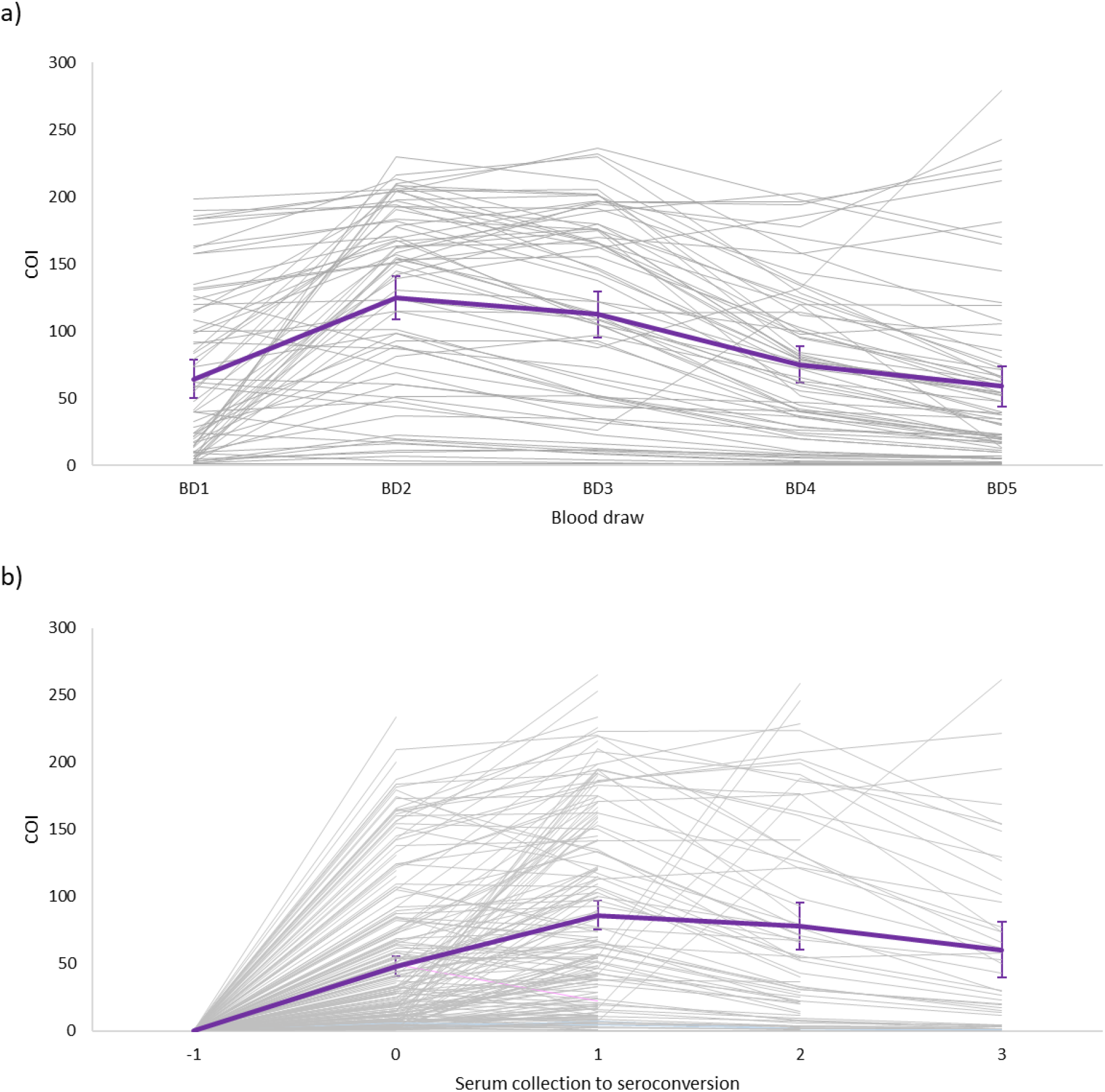
Cut-off index (COI) on Roche Elecsys® Anti-SARS-CoV-2 assay for individuals with blood draw 1 to 5 (BD1 to BD5) samples who a) were seropositive at baseline b) seroconverted during blood draw 2-5, July 2020 – April 2021, South Africa. Mean COI with 95% confidence interval in purple line. COI values in b aligned to first draw prior to seroconversion.

## Discussion

We assessed SARS-CoV-2 seroprevalence in 1,211 individuals living in two diverse communities in South Africa and show that laboratory-confirmed cases reported from study districts greatly underestimate the true burden of SARS-CoV-2 infections. At baseline, seroprevalence was 1% and 15%, increasing to 7% and 27% after the first wave, and by March 2021. Following the second epidemic wave, seroprevalence was 26% and 41% in the rural and urban communities, respectively. The highest seroprevalence was 59% in adults aged 35-59 years in the urban community, and the lowest was 18% in rural community children <5 years. During the second wave, compared to the first wave, the rural site was more affected, and infections in the second wave more likely affected children aged 5-12 years and adults ≥60 years. We observed no differences in seroprevalence by HIV status. In the urban community, the IFR was higher in the second wave (0.36-0.51%), compared to the first (0.13-0.17%), even though numbers of infections were lower, suggesting possible increased severity associated with the emergence of novel variant 501Y.V2. Most individuals who seroconverted maintained detectable SARS-CoV-2 antibodies in subsequent serum samples.

Low seropositivity was observed at the rural site at baseline, and the seroprevalence remained low until blood draw 3, only reaching seroprevalence of 7% after the first wave of infections, which was considerably lower than the seroprevalence of 27% at the urban site at the same time. This could possibly be related to the relatively isolated location and lower population density in the rural community compared to more densely populated urban community. The seroprevalence in the rural site increased to 26% at blood draw 5, which was after the second wave of infections within the district. This could have been due to possible increased transmissibility of the 501Y.V2 lineage that was circulating in the second wave,^20^ as well as additional transmission networks in the community during the December holiday period when large scale urban-to-rural migration takes place as people return home for the yearend holidays. The urban site had fewer seroconversions in the second wave compared to the first, which may be due to existing immunity among individuals in the community after the first wave. As seen in previous studies,^9^ adults had the highest seroprevalence levels, although there was still a relatively high seroprevalence of 18% and 28% in children <5 years at the rural and urban community respectively.

There are few data available on seroprevalence in Africa, most studies have focused on specialised groups. A study conducted among blood donors in South Africa during the second wave of the pandemic found a seroprevalence of 32% to 63%, in five provinces of South Africa that have both rural and urban communities.^21^ In our study, enrolling a random sample of community members, we observed a seroprevalence in adults ranging between 25-37% in rural households, and 35-59% in the urban households, suggesting that seroprevalence is heterogeneous between communities. Seroprevalence data from sub-Saharan Africa are also limited. In Kenya the seroprevalence in blood donors during the country’s first wave of infections was 4%, and was also higher in urban communities.^22^ In a population-level household sero-survey in Zambia during their first wave of infections, 11% of individuals had evidence of SARS-CoV-2 infection.^23^ After the first wave, we estimated a higher seroprevalence in the rural (7%) and in the urban communities (27%) compared to Kenya^22^ and a higher seroprevalence in the urban community than in Zambia.^22,23^

Based on our estimates, only 4-6% of cases were laboratory confirmed. Our study suggests substantially higher prevalence of infection ascertained through serology and that the differences may have been greater in the urban community than the rural community; however, more extensive studies are needed to assess whether this is consistent in other areas. Compared to the urban community, the rural community had a higher proportion of SARS-CoV-2 infections reported to the NMCSS (4-6% vs 4%) and less than half the rate of hospitalisation (0.8% vs 2-3%). These may be due to differences in referral and testing policies, health-seeking behaviour, access to care and differences in circulating lineages within these districts.

A study comparing the severity of the first and second waves of infections in South Africa in hospitalised patients found a higher mortality rate in the second wave, compared to the first, even controlling for increased pressure on health services which was also associated with increased mortality.^24^ At the urban site, the IFR was significantly higher in the second wave (0.43-0.50%) compared to the first (0.13-0.20%), although no differences were observed in IHR between the two waves. The lower overall number of infections in the second wave in this site means that our finding of increased mortality is unlikely to be related to pressure on health services. The increased severity of the second wave may be related to increased severity of the 501Y.V2/B.1.135 variant, but further studies are needed to confirm this. The excess death IFR during the first wave in the urban site was smaller than the in-hospital IFR. This may be due to uncertainty on the process for excess death estimation, or that the 85% contribution of COVID-19 to excess deaths was an underestimation within the province. However, the in-hospital IFR followed the same trend of increase between wave 1 and 2 (0.13% to 0.51%).

Considering the high seroprevalence observed, the age standardised IHR (wave 1: 0.8% rural, 2% urban, wave 2: 0.8% rural, 2.58% urban) and IFR (wave 1: 0.2-0.5% rural, 0.1-0.2% urban, wave 2: 0.2-0.4% rural, 0.4-0.5% urban) were lower compared to the non-age standardised estimates from the USA, where the IHR and IFR was estimated as 2% and 1% respectively,^25^ and from Italy where IFR was estimated as 0.9%.^26^ Our first wave in-hospital IFR estimates (0.13% rural, 0.17% urban) were more similar to the age-adjusted 0.15% reported from India for the first wave SARS-CoV-2-confirmed deaths,^27^ and our first wave excess death IFR was higher in the rural (0.51%) and lower in the urban (0.13%) community compared to the age-adjusted 0.28% IFR excess deaths reported from Brazil during their first wave of infections.^28^

We did not observe a difference in SARS-CoV-2 seroprevalence in HIV-infected and –uninfected individuals at either site. Due to HIV causing immune suppression, there is a concern that HIV-infected individuals may be more susceptible to SARS-CoV-2 infection.^10,11^ A meta-analysis including data from North America, Africa, Europe, and Asia found a 24% higher risk of symptomatic SARS-CoV-2 infection in HIV-infected individuals, albeit with high heterogeneity between countries,^11^ whereas a case-control study from the United States of America found a lower SARS-CoV-2 seroprevalence in HIV-infected individuals. Although HIV infection may not increase susceptibility to infection, it has been demonstrated to be a risk factor for developing severe COVID-19 and death following infection.^10,29^

Our study is limited by a relatively small sample size, reducing the power for accurate seroprevalence estimates in small age strata; and including only two geographic sites, and therefore may not be representative of other districts and provinces in South Africa. The seroprevalence reported here may be an under-estimate of population-level infection rates as not everyone infected with SARS-CoV-2 develops antibodies. ICR, IHR and IFR formed part of an ecological analysis which is inherently prone to biases. Excess deaths in the first wave may be underestimated since the reporting period only stared in June, and were reported at provincial-level which may be different to within the district. Similarly, transmission dynamics within our cohort may not be similar to the district which formed the comparison point for our case, hospitalisation and in-hospital deaths. The ELISA utilised to detect SARS-CoV-2 antibodies was qualitative, and not suited for quantitative analysis of antibody levels. Ongoing follow-up of this cohort will track future infections and monitor antibody waning, and compare these data to laboratory confirmed infections and symptoms from twice weekly follow-up. A strength of our study is the collection of samples from prospectively followed up individuals from randomly selected households within the study communities and inclusion of individuals of all ages. To our knowledge, this is the first seroprevalence data from a cohort in South Africa, which provides the advantage of serial comparisons of antibody responses in relation to reported laboratory-confirmed SARS-CoV-2 infections within the community through two successive SARS-CoV-2 waves.

We estimate that approximately 95% of SARS-CoV-2 infections in these two communities were not laboratory-confirmed and reported to the national surveillance system, which has major implications for contact tracing and isolation and other measures to contain infection. We observed heterogeneity between seroprevalence estimates based on pandemic wave, community and age group indicating the need for ongoing studies with inclusion of diverse settings.

## Data Availability

The investigators welcome enquiries about possible collaborations and requests for access to the dataset. Data will be shared after approval of a proposal and with a signed data access agreement. Investigators interested in more details about this study, or in accessing these resources, should contact the corresponding author.

## Acknowledgements

All individuals participating in the study, field teams for their hard work and dedication to the study, especially Dr Tumelo Moloantoa, Dr Kgaugelo Kgasago, Nompumelelo Yende and Sizzy Ngobeni for coordination of site activities, Charlotte Yaldwyn and Shingirai Chipungu for processing of collected bloods, the CRDM laboratory team, Doreen Janse van Rensburg for managing logistics surrounding testing of collected bloods and result extraction, Nevashan Govender, Genevie Ntshoe, Andronica Moipone Shonhiwa, Darren Muganhiri, Itumeleng Matiea, Eva Mathatha, Fhatuwani Gavhi, Teresa Mashudu Lamola, Matimba Makhubele, Mmaborwa Matjokotja, Simbulele Mdleleni, Masingita Makhubela from the national SARS-CoV-2 NICD surveillance team, Lincoln Darwin, Fazil Mckenna, Dr Trevor Graham Bell, Ndivhuwo Munava, Muzammil Raza Bano and Jimmy Khosa from NICD IT, Prof Rob Dorrington from the Centre for Actuarial Research for providing wave-specific excess death estimates, Ben Cowling, Kanta Subbarao, Juliet Pulliam, Melissa Rolfes and Mosa Moshabela from the PHIRST-C scientific committee and Andrew Whitelaw, June Fabian, Jennifer Verani, Lindiwe Qwabe, Banele Faku and Saheen Methar from the PHIRST-C safety committee.

## Disclaimer

The findings and conclusions in this report are those of the authors and do not necessarily represent the official position of the CDC.

