## Appendix for "Longitudinal SARS-CoV-2 seroprevalence in a rural and urban community household cohort in South Africa, during the first and second waves July 2020-March 2021"

### Methods

*Study population*

PHIRST-C (A Prospective Household study of SARS-CoV-2, Influenza, and Respiratory Syncytial virus community burden, Transmission dynamics and viral interaction in South Africa) is a continuation from PHIRST (A Prospective Household observational cohort study of Influenza, Respiratory Syncytial virus and other respiratory pathogens community burden and Transmission dynamics in South Africa).[1] PHIRST-C is conducted in two communities with established influenza-like illness and pneumonia surveillance sites (Appendix figure 1).

South Africa consists of nine provinces, which are divided into 52 districts, which form the second level of administration. Districts are further divided into municipalities. The rural site is located in the Bushbuckridge Municipality, Ehlanzeni District, Mpumalanga Province and forms part of a health and socio-demographic surveillance site (HDSS) at the Medical Research Council (MRC)/University of Witwatersrand Rural Public Health and Health Transitions Research Unit, Agincourt. During PHIRST, two of the 29 villages within the HDSS were selected according to convenience (proximity and burden of other studies within the site) each year between 2016-2018. A random selection of ~50 households with a household size of 3 or more were approached for enrolment. The urban site is located in the Jouberton Township, Matlosana Municipality, Dr Kenneth Kaunda District, North West Province. A list of 450 global positioning system (GPS) coordinates were generated using Google Earth. Study staff approached the nearest house within 30 meters of the coordinate point for enrolment. Approximately 50 households were enrolled each year from 2016-2018. At both sites, households were eligible if they consisted of three or more household members (sharing at least four meals a week). All household members were approached for inclusion in the study, and households were eligible for inclusion if >80% of members consented to be enrolled.

For PHIRST-C, all households who participated in PHIRST from the urban site, and households from the 2017 and 2018 cohort in the rural site were approached for enrolment. To appendix the sample size, additional households at each site were approached using the same methods as for PHIRST. Villages in the rural site were restricted to those four used in 2017 and 2018 of PHIRST. Informed consent was obtained from participating adults or a parent/guardian for children aged <18 years. In addition to parent/guardian consent, assent was also obtained from children aged 7-17 years.

The Bushbuckridge Municipality within the Ehlanzeni District is considered as predominantly rural, with main industries being agriculture and tourism.[2] The Ehlanzeni District has a population of 1,828,738, with sixty percent of the district's population being >18 years of age (Appendix Table 1). The City of Matlosana Municipality is considered to be 88.2% urban, and is located in the Dr Kenneth Kaunda District, which has a population of 797,715.[3] Sixty-five percent of the district population is aged >18 years (Appendix Table 1). At both sites, the percentage of children, females, and unemployed individuals were higher in the cohort than in the district (Appendix Table 1).

*Data sources*

- Population denominators [4]

  Population numbers for each district, by age, were obtained from the StatsSA 2020 mid-year population estimates for 2020.

- Notifiable Medical Conditions Surveillance System (NMCSS) [5]

  Reverse transcription polymerase chain reaction (RT-PCR) testing in South Africa to detect SARS-CoV-2 RNA started on 28 January 2020. Rapid SARS-CoV-2 antigen testing was implemented in November 2020. All laboratories in the private and public sector performing SARS-CoV-2 tests automatically feed testing data from their lab information systems to the NMCSS, from where daily and weekly reporting on national, provincial and district SARS-CoV-2 infections are performed. No serology tests are used for this reporting. Results from antigen tests may be underestimated in the NMCSS. The number of cases reported to the NMCSS in the Ehlanzeni District (rural community) and Dr Kenneth Kaunda District (urban community) was used as an estimate of reported, laboratory-confirmed SARS-CoV-2 infections. Wave 1: 3 March – 21 November 2020, wave 2: 22 November 2020 – 27 March 2021.

- COVID-19 National Hospital Surveillance (DATCOV) [6]

  DATCOV is a national hospital surveillance system to which all hospitals where COVID-19 admissions occurred in the public and private sector in South Africa report to. The case definition includes any person admitted with a positive RT-PCR test for SARS-CoV-2. In-hospital outcome was available for all patients. The total number of hospitalisations and in-hospital deaths in each district reported to DATCOV between 3 March – 21 November 2020 (wave 1) and 22 November 2020 – 27 March 2021 (wave 2) was used to calculate in the IHR and minimum IFR, respectively.

- South African Medical Research Council (SAMRC) report on weekly deaths [7]

  The Burden of Disease Unit at the SAMRC produces a weekly report on excess deaths in South Africa. Deaths in South Africa are registered on the Department of Home Affair's National Population Register and includes all citizens with a South African identification number. The number of excess deaths were defined as the number of all-cause deaths in that week minus the number of deaths expected for the week based on 2014-2019 trends. Excess death estimates are adjusted for incomplete reporting, and death estimates in provinces are age-standardised to the national population. Eighty-five percent of the reported excess deaths were used to calculate the maximum IHR. In both provinces, excess deaths were only available starting 21 June (urban) and 28 June 2020 (rural), and not from March as for infections and hospitalisations, therefore the period for wave 1 in the rural community was defined as 28 June – 21 November 2020, and as 21 June – 21 November 2020 in urban communities for wave 1. For both communities wave 2 was defined as 22 November 2021 – 26 March 2021. The wave-specific excess death rates were kindly provided by Prof. Rob Dorrington.

### Appendix tables

**Appendix Table 1**. Comparison of characteristics between district (Ehlanzeni and Dr Kenneth Kaunda Districts)[4] and PHIRST-C cohort population, 2020, South Africa.

| | Ehlanzeni District | Rural community cohort | p-value | Dr Kenneth Kaunda Districts | Urban community cohort | p-value |
|---|---|---|---|---|---|---|
| **Age group (years)** | | | | | | |
| **<5** | 203727/1828739 (11%) | 99/668 (15%) | <0.01 | 75574/797716 (9%) | 56/598 (9%) | 0.93 |
| **5-12** | 319208/1828739 (17%) | 210/668 (31%) | <0.01 | 121145/797716 (15%) | 132/598 (22%) | <0.01 |
| **13-18** | 207344/1828739 (11%) | 89/668 (13%) | 0.11 | 82582/797716 (10%) | 82/598 (14%) | 0.01 |
| **19-34** | 491914/1828739 (27%) | 108/668 (16%) | <0.01 | 212152/797716 (27%) | 120/598 (20%) | <0.01 |
| **35-59** | 462982/1828739 (25%) | 89/668 (13%) | <0.01 | 237146/797716 (30%) | 122/598 (20%) | <0.01 |
| **≥60** | 143564/1828739 (8%) | 46/668 (7%) | 0.35 | 69117/797716 (9%) | 58/598 (10%) | 0.37 |
| **Female sex** | 961384/1828739 (53%) | 427/668 (64%) | <0.01 | 405734/797716 (51%) | 336/598 (56%) | 0.01 |
| **Unemployed** | 187428/530865 (35%) | 198/270 (73%) | <0.01 | 76654/257457 (30%) | 202/315 (64%) | <0.01 |

**Appendix Table 2**. Number of sera collected (n) from consenting PHIRST-C participants (N) and percentage sampled during blood draws (BD) in the rural and urban community by blood draw, site, age group, March 2020 – March 2021, South Africa.
Baseline (BD1) 20 July – 17 September 2020, second draw (BD2) 21 September – 10 October 2020, third draw (BD3) 23 November – 12 December 2020, fourth draw (BD4) 25 January – 20 February 2021, fifth draw (BD5) 22 March – 11 April 2021.

| | **Rural n/N (%)** | | | | | | |
|---|---|---|---|---|---|---|---|
| | **B1** | **B2** | **B3** | **B4** | **B5** | **B1-5** | **B3,5** |
| **<5 years** | 25/102 (25) | 49/102 (48) | 78/102 (76) | 87/102 (85) | 89/102 (87) | 17/102 (17) | 76/102 (75) |
| **5-12 years** | 148/215 (69) | 169/215 (79) | 195/215 (91) | 202/215 (94) | 201/215 (93) | 136/215 (63) | 193/215 (90) |
| **13-18 years** | 69/94 (73) | 72/94 (77) | 80/94 (85) | 82/94 (87) | 78/94 (83) | 61/94 (65) | 76/94 (81) |
| **19-34 years** | 87/117 (74) | 88/117 (75) | 91/117 (78) | 91/117 (78) | 95/117 (81) | 62/117 (53) | 89/117 (76) |
| **35-59 years** | 76/91 (84) | 77/91 (85) | 80/91 (88) | 81/91 (89) | 82/91 (90) | 66/91 (73) | 78/91 (86) |
| **≥60 years** | 40/49 (82) | 39/49 (80) | 44/49 (90) | 45/49 (92) | 42/49 (86) | 35/49 (71) | 41/49 (84) |
| **All ages** | 445/668 (67) | 494/668 (74) | 568/668 (85) | 588/668 (88) | 587/668 (88) | 377/668 (56) | 553/668 (83) |
| | **Urban n/N (%)** | | | | | | |
| | **B1** | **B2** | **B3** | **B4** | **B5** | **B1-5** | **B3,5** |
| **<5 years** | 45/57 (79) | 50/57 (88) | 50/57 (88) | 44/57 (77) | 48/57 (84) | 33/57 (58) | 45/57 (79) |
| **5-12 years** | 116/135 (86) | 116/135 (86) | 125/135 (93) | 123/135 (91) | 122/135 (90) | 103/135 (76) | 120/135 (89) |
| **13-18 years** | 75/84 (89) | 75/84 (89) | 81/84 (96) | 77/84 (92) | 78/84 (93) | 69/84 (82) | 78/84 (93) |
| **19-34 years** | 102/130 (78) | 98/130 (75) | 100/130 (77) | 99/130 (76) | 98/130 (75) | 77/130 (59) | 92/130 (71) |
| **35-59 years** | 110/130 (85) | 110/130 (85) | 117/130 (90) | 112/130 (86) | 111/130 (85) | 98/130 (75) | 111/130 (85) |
| **≥60 years** | 57/62 (92) | 55/62 (89) | 56/62 (90) | 55/62 (89) | 53/62 (85) | 51/62 (82) | 53/62 (85) |
| **All ages** | 505/598 (84) | 504/598 (84) | 529/598 (88) | 510/598 (85) | 510/598 (85) | 431/598 (72) | 499/598 (83) |

**Appendix Table 3**. Individuals seropositive for SARS-CoV-2 antibodies with adjusted seroprevalence and 95% creditable intervals by blood draw, site and age, July 2020 – April 2021, South Africa. Baseline (BD1) 20 July – 17 September 2020, second draw (BD2) 21 September – 10 October 2020, third draw (BD3) 23 November – 12 December 2020, fourth draw (BD4) 25 January – 20 February 2021, fifth draw (BD5) 22 March – 11 April 2021.

| | | BD1 | BD2 | BD3 | BD4 | BD5 |
|---|---|---|---|---|---|---|
| | **<5 years** | 0/25 (4, 0-13) | 1/49 (4, 0-13) | 2/78 (4, 1-9) | 14/87 (17, 10-25) | 15/89 (18, 10-26) |
| | **5-12 years** | 2/148 (2, 0-5) | 5/169 (3, 0-5) | 7/195 (4, 2-7) | 29/202 (15, 10-20) | 39/201 (20, 14-25) |
| | **13-18 years** | 1/69 (3, 0-7) | 5/72 (8, 0-7) | 8/80 (11, 5-18) | 20/82 (25, 16-35) | 24/78 (31, 22-42) |
| **Rural** | **19-34 years** | 0/87 (1, 0-4) | 5/88 (7, 0-4) | 13/91 (15, 8-23) | 33/91 (37, 27-47) | 35/95 (37, 28-47) |
| | **35-59 years** | 1/76 (2, 0-7) | 5/77 (7, 0-7) | 7/80 (10, 4-17) | 23/81 (29, 20-39) | 27/82 (33, 23-44) |
| | **≥60 years** | 1/40 (5, 0-13) | 4/39 (12, 0-13) | 4/44 (11, 3-21) | 8/45 (19, 9-32) | 10/42 (25, 13-38) |
| | **All ages** | 5/445 (1, 0-2) | 25/494 (5, 0-2) | 41/568 (7, 5-9) | 127/588 (22, 18-25) | 150/587 (26, 22-29) |
| | **<5 years** | 2/45 (6, 1-15) | 5/50 (11, 4-21) | 7/50 (15, 7-26) | 8/44 (20, 9-32) | 13/48 (28, 17-41) |
| | **5-12 years** | 11/116 (10, 5-16) | 17/116 (15, 9-22) | 24/125 (20, 13-27) | 32/123 (26, 19-34) | 38/122 (31, 23-40) |
| | **13-18 years** | 14/75 (19, 11-29) | 19/75 (26, 17-36) | 30/81 (37, 27-48) | 35/77 (46, 35-56) | 41/78 (53, 42-64) |
| **Urban** | **19-34 years** | 10/102 (10, 5-17) | 17/98 (18, 11-26) | 23/100 (23, 16-32) | 29/99 (30, 21-39) | 34/98 (35, 26-44) |
| | **35-59 years** | 33/110 (30, 22-39) | 45/110 (41, 32-50) | 52/117 (45, 36-54) | 61/112 (55, 45-64) | 65/111 (59, 49-68) |
| | **≥60 years** | 3/57 (7, 2-14) | 6/55 (12, 5-22) | 7/56 (14, 6-24) | 16/55 (30, 19-42) | 18/53 (35, 22-47) |
| | **All ages** | 73/505 (15, 12-18) | 109/ (22, 18-25) | 143/529 (27, 23-31) | 181/510 (36, 32-40) | 209/510 (41, 37-45) |

**Appendix Table 4**. SARS-CoV-2 seroprevalence from the fifth blood collection (22 March – 11 April 2021), by site, age group and HIV status, South Africa.

| | Rural | | | Urban | | |
|---|---|---|---|---|---|---|
| | **HIV-uninfected** | **HIV-infected** | **p-value** | **HIV-uninfected** | **HIV-infected** | **p-value** |
| **<5 years** | 15/87 (17) | 0/1 (0) | 0.65 | 12/46 (26) | 0/1 (0) | 0.55 |
| **5-12 years** | 37/195 (19) | 0/2 (0) | 0.49 | 35/116 (30) | 1/3 (33) | 0.91 |
| **13-18 years** | 21/66 (32) | 0/3 (0) | 0.24 | 38/73 (52) | 3/4 (75) | 0.37 |
| **19-34 years** | 22/65 (34) | 10/23 (43) | 0.41 | 26/73 (36) | 7/19 (37) | 0.92 |
| **35-59 years** | 12/36 (33) | 12/39 (31) | 0.81 | 37/61 (61) | 28/47 (60) | 0.91 |
| **≥60 years** | 6/27 (22) | 4/10 (40) | 0.28 | 17/46 (37) | 1/7 (14) | 0.24 |
| **All ages** | 113/476 (24) | 26/78 (33) | 0.07 | 165/415 (40) | 40/81 (49) | 0.11 |

**Appendix figures**

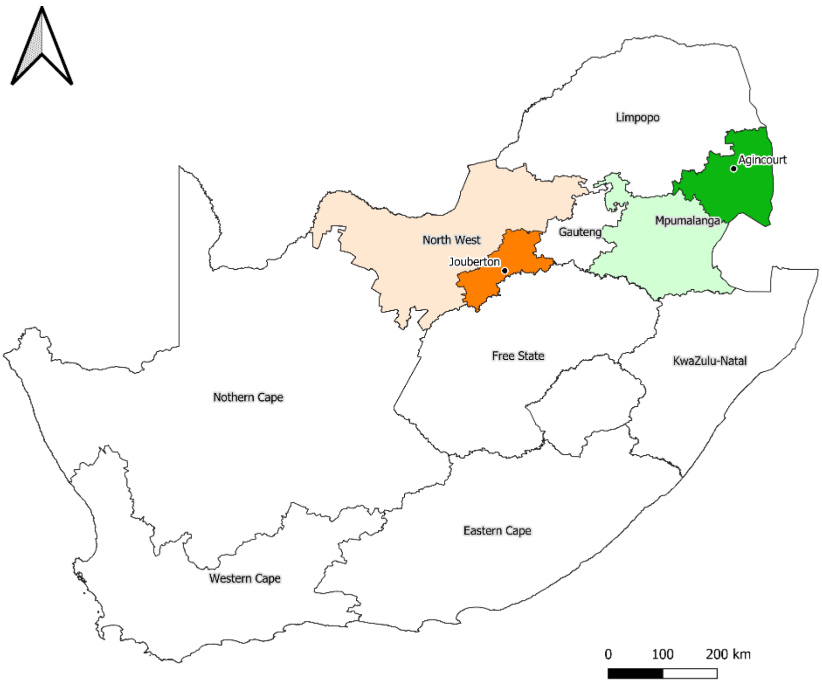

**Appendix Figure 1**. Location of study sites PHIRST-C, South Africa.

The rural site is located in Agincourt, Ehlanzeni District (dark green), Mpumalanga Province (light green) and the urban site in Jouberton, Dr Kenneth Kaunda District (dark orange), North West Province (light orange).

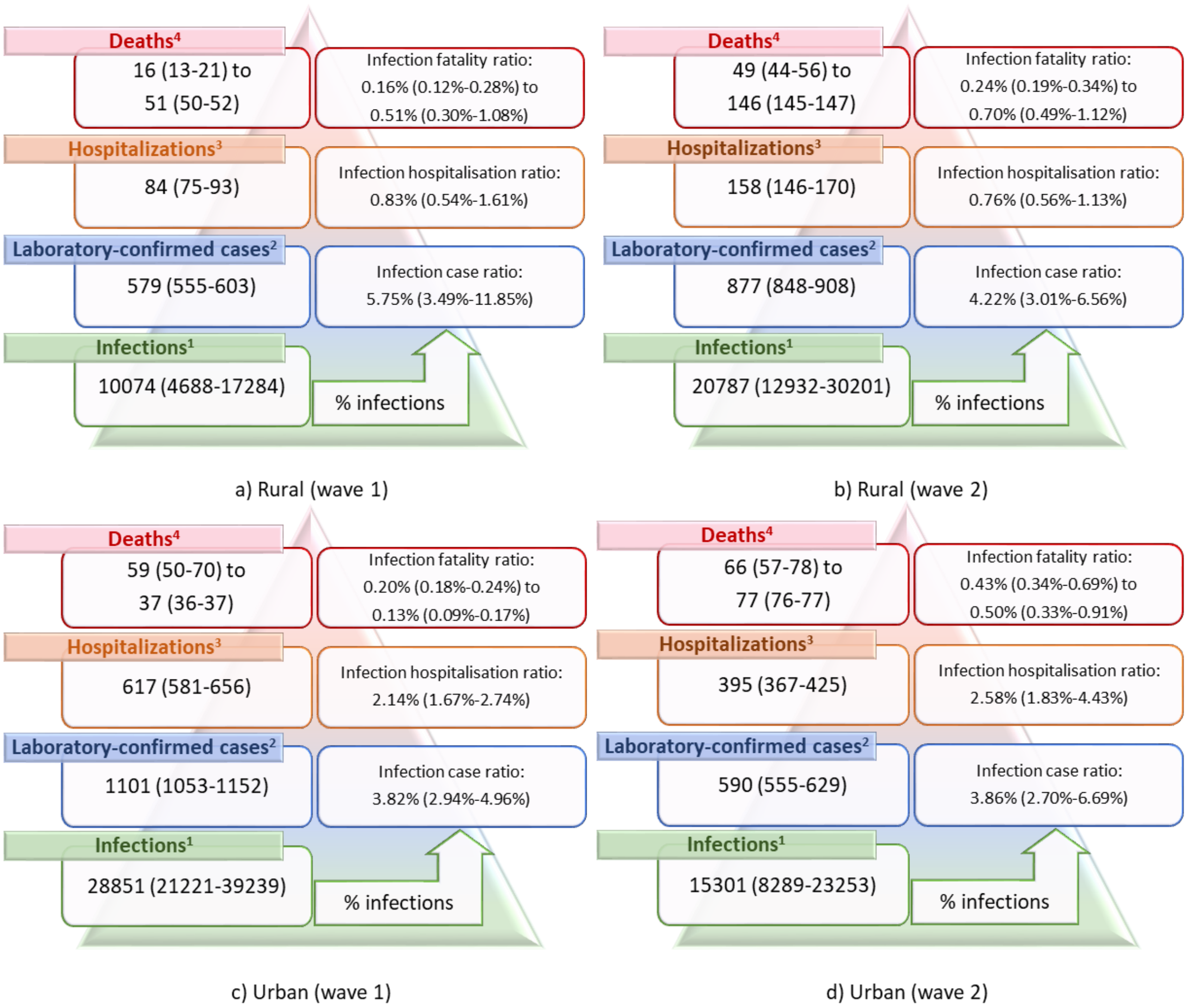

**Appendix Figure 2.** WHO population age-standardised SARS-CoV-2 infection, diagnosis, hospitalisation and deaths per 100,000 population in a rural community during a) wave 1 and b) wave 2, and urban community during c) wave 1 and d) wave 2 of infections, South Africa, March 2020 - March 2021.

[1] Based on seroprevalence at third/fifth blood draw. [2] Laboratory confirmed SARS-CoV-2 cases reported from districts from national SARS-CoV-2 surveillance (NMCSS, wave 1: 3 March – 21 November 2020, wave 2: 22 November 2020 – 27 March 2021). [3] Hospitalisations in the district based on COVID-19 Sentinel Hospital Surveillance (DATCOV, wave 1: 5 March – 21 November 2020, wave 2: 22 November 2020 – 27 March 2021). [4] Minimum estimate: in-hospitalisation deaths in districts based on COVID-19 Sentinel Hospital Surveillance report (DATCOV, wave 1: 5 March – 21 November 2020, wave 2: 22 November 2020 – 27 March 2021), maximum estimate: provincial excess deaths reported by South African Medical Research Council (rural wave 1: 28 June – 21 November 2020, urban wave 1: 21 June – 21 November 2020, wave 2: 22 November 2021 – 26 March 2021). Wave 1: 1 March – 21 November 2020), wave 2 22 November 2020 – 27 March 2021. Values in bracket refer to 95% credible interval for infections and 95% confidence interval for all other estimates

**The PHIRST-C Group**

| Name | Affiliation |
|---|---|
| Amelia Buys | Centre for Respiratory Diseases and Meningitis, National Institute for Communicable Diseases of the National Health Laboratory Service, Johannesburg, South Africa. |
| Anne von Gottberg | Centre for Respiratory Diseases and Meningitis, National Institute for Communicable Diseases of the National Health Laboratory Service, Johannesburg, South Africa. School of Pathology, Faculty of Health Sciences, University of the Witwatersrand, Johannesburg, South Africa. |
| Cheryl Cohen | Centre for Respiratory Diseases and Meningitis, National Institute for Communicable Diseases of the National Health Laboratory Service, Johannesburg, South Africa. School of Public Health, Faculty of Health Sciences, University of the Witwatersrand, Johannesburg, South Africa. |
| F. Xavier Gómez-Olivé | MRC/Wits Rural Public Health and Health Transitions Research Unit (Agincourt), Faculty of Health Sciences, School of Public Health, University of the Witwatersrand, Johannesburg, South Africa. |
| Floidy Wafawanaka | MRC/Wits Rural Public Health and Health Transitions Research Unit (Agincourt), Faculty of Health Sciences, School of Public Health, University of the Witwatersrand, Johannesburg, South Africa. |
| Jackie Kleynhans | Centre for Respiratory Diseases and Meningitis, National Institute for Communicable Diseases of the National Health Laboratory Service, Johannesburg, South Africa. School of Public Health, Faculty of Health Sciences, University of the Witwatersrand, Johannesburg, South Africa. |
| Jacques du Toit | MRC/Wits Rural Public Health and Health Transitions Research Unit (Agincourt), Faculty of Health Sciences, School of Public Health, University of the Witwatersrand, Johannesburg, South Africa. |
| Jinal N. Bhiman | Centre for Respiratory Diseases and Meningitis, National Institute for Communicable Diseases of the National Health Laboratory Service, Johannesburg, South Africa. School of Pathology, Faculty of Health Sciences, University of the Witwatersrand, Johannesburg, South Africa. |
| Jocelyn Moyes | Centre for Respiratory Diseases and Meningitis, National Institute for Communicable Diseases of the National Health Laboratory Service, Johannesburg, South Africa. School of Public Health, Faculty of Health Sciences, University of the Witwatersrand, Johannesburg, South Africa. |
| Kathleen Kahn | MRC/Wits Rural Public Health and Health Transitions Research Unit (Agincourt), Faculty of Health Sciences, School of Public Health, University of the Witwatersrand, Johannesburg, South Africa. |
| Kgaugelo Patricia Kgasago | Perinatal HIV Research Unit (PHRU), University of the Witwatersrand, Johannesburg, South Africa. |
| Limakatso Lebina | Perinatal HIV Research Unit (PHRU), University of the Witwatersrand, Johannesburg, South Africa. |
| Linda de Gouveia | Centre for Respiratory Diseases and Meningitis, National Institute for Communicable Diseases of the National Health Laboratory Service, Johannesburg, South Africa. |
| Maimuna Carrim | Centre for Respiratory Diseases and Meningitis, National Institute for Communicable Diseases of the National Health Laboratory Service, Johannesburg, South Africa. |

| | |
|---|---|
| Meredith L. McMorrow | Influenza Division, Centers for Disease Control and Prevention, Atlanta, Georgia, United States of America. Influenza Program, Centers for Disease Control and Prevention, Pretoria, South Africa. |
| Mignon du Plessis | Centre for Respiratory Diseases and Meningitis, National Institute for Communicable Diseases of the National Health Laboratory Service, Johannesburg, South Africa. |
| Neil A. Martinson | Perinatal HIV Research Unit (PHRU), University of the Witwatersrand, Johannesburg, South Africa. Johns Hopkins University Center for TB Research, Baltimore, Maryland, United States of America. |
| Nicole Wolter | Centre for Respiratory Diseases and Meningitis, National Institute for Communicable Diseases of the National Health Laboratory Service, Johannesburg, South Africa. School of Pathology, Faculty of Health Sciences, University of the Witwatersrand, Johannesburg, South Africa. |
| Retshidisitswe Kotane | Centre for Respiratory Diseases and Meningitis, National Institute for Communicable Diseases of the National Health Laboratory Service, Johannesburg, South Africa. School of Pathology, Faculty of Health Sciences, University of the Witwatersrand, Johannesburg, South Africa. |
| Stefano Tempia | Influenza Division, Centers for Disease Control and Prevention, Atlanta, Georgia, United States of America. Influenza Program, Centers for Disease Control and Prevention, Pretoria, South Africa. School of Public Health, Faculty of Health Sciences, University of the Witwatersrand, Johannesburg, South Africa. MassGenics, Atlanta, Georgia, United States of America. |
| Stephen Tollman | MRC/Wits Rural Public Health and Health Transitions Research Unit (Agincourt), School of Public Health, Faculty of Health Science, University of the Witwatersrand. |
| Tumelo Moloantoa | Perinatal HIV Research Unit (PHRU), University of the Witwatersrand, Johannesburg, South Africa. |
